# Learning latent profiles via cognitive growth charting in psychosis: design and rationale for the PRECOGNITION project

**DOI:** 10.1101/2024.11.26.24317909

**Authors:** Andre F. Marquand, Barbora Rehák Bučková, Giulia Cattaranusi, Camilla Flaaten, Cecilie Busch, Cecilie K. Lemvigh, Veenu Gupta, Charlotte Fraza, Lars T. Westlye, Ole A. Andreassen, Jaroslav Hlinka, Bjørn H. Ebdrup, David Shiers, Torill Ueland, Paola Dazzan

## Abstract

Cognitive impairments are a core feature of psychosis that are often evident before illness onset and have substantial impact on both clinical and real-world functional outcomes. Therefore, these are an excellent target for stratification and early detection in order to facilitate early intervention. While many studies have aimed to characterise the effects of cognition at the group level and others have aimed to detect individual differences by referencing subjects against existing norms, these studies have limited generalisability across clinical populations, demographic backgrounds and instruments and also do not fully account for the inter-individual heterogeneity inherent in psychosis. Here, we outline the rationale, design and analysis plan for the PRECOGNITION project which aims to address these challenges. This project is a collaboration between partners in five European countries, that aims to (i) translate normative modelling approaches that have been pioneered in brain imaging to psychosis data, to yield ‘cognitive growth charts’ for longitudinal tracking and prediction at the individual level (ii) develop machine learning models for harmonising and stratifying cohorts on the basis of these models; (iii) provide comprehensive reference models that provide broad sociodemographic coverage including different languages and distinct norms for individuals with psychosis and unaffected individuals and (iv) stratification models and predictions for functional outcome measures in psychosis cohorts. Crucially, this project will be guided throughout by experts with lived experience.

## Introduction

Cognitive impairments, such as impairments in memory, reasoning and concentration, are a core feature of psychosis. They are often evident before illness onset^1–5^ and have substantial impact on functional outcomes and activities of daily living^6^. This suggests that cognitive functions should be a priority target for developing early detection and stratification approaches to facilitate early intervention. However, while cognitive alterations may have high sensitivity early in the course of the illness, they have low specificity in that impairments are present across several cognitive domains^2^ and are also evident early in the course of a range of common behavioural and mental disorders. Indeed, a recent meta-analytic report^4^ provided evidence that group level impairments are evident across all cognitive domains in first episode psychosis, bipolar disorder and depression. Moreover, differentiation between disorders is principally evident in the magnitude of the effects reported^4^ rather than in the nature of the effects, although some differences exist^4^. This suggests that precise quantification of the magnitude, timing and progression of cognitive impairments are essential in order to parse the heterogeneity underlying psychosis.

An additional problem is that studies to date have overwhelmingly focussed on group level analyses, which mask considerable inter-individual variation that may have clinical relevance. This limitation is a major barrier in our ability to predict the onset and course of illness and to the development of personalized early interventions. The heterogeneity of mental disorders is well recognised in theoretical models^7–11^ and the importance of modelling individual differences is widely acknowledged^10–13^. However, this theoretical recognition is not reflected in the typical ‘case-control’ paradigm often used to study psychiatric disorders: nearly all approaches used in practice are implicitly rooted in the ‘case-control’ paradigm which assumes clinical groups are homogeneous and well-defined (patient/control, responder/remitter).

While this has helped to partly understand the mechanisms of mental disorders, this assumption of clean distinctions between clinical groups is often unrealistic and focusing on group effects masks inter-individual variation that may be crucial for individual prediction. For example, the clinical group may be diffuse and heterogeneous due to comorbidity and to a convergence of different pathophysiological pathways on the same symptoms. In line with this, there has been considerable effort invested in finding subgroups of patients based on cognition or symptoms over the last 50 years and many putative stratifications have been proposed^14–20^, yet none have advanced beyond proof-of-concept. We have suggested that one of the main reasons is that clustering algorithms always yield a result, regardless of whether clearly defined clusters are evident in the data, resulting in ill-defined and poorly replicated subtypes.^21,22^

Whilst clinical neuropsychology has traditionally adopted a single subject perspective by referencing subjects against group-level norms, these approaches also have limitations. For example, in a research context, most studies also apply group level analyses to detect average differences between cohorts based on normed data. Moreover, existing norms are typically generated for specific tests or batteries and based on a single population. These have limited generalisability across different tests, more demographically representative populations and across languages. Also, existing approaches do not fully capture the potentially complex shape of different test scores, which often have non-linear, non-Gaussian and heteroskedastic effects across the lifespan. Finally, it is questionable whether the same norms should be applied to healthy individuals and to individuals with psychosis. Indeed, we have shown that the variability amongst individuals with psychosis is greater than within the healthy population for most cognitive functions and norms estimated on healthy populations are therefore suboptimal for understanding inter-individual differences in psychosis (e.g. due to the presence of floor or ceiling effects).^23^ We have also provided proof of concept evidence that the approach we propose here can help to make progress.^24^

The Wellcome-Trust funded <u>PRECOGNITION</u> project aims to address these problems, by applying the normative modelling approach developed in brain imaging to cognitive data. The normative modelling framework generalises the notion of pediatric growth charts aiming to model centiles of variation in the population as a function of clinically relevant covariates, whilst providing statistical inferences at the level of the individual person.^21,25^ It has proved to be especially useful in understanding inter-individual variability in psychosis.^26–30^ More specifically, this project aims to: (i) apply normative modelling to cognitive data, to yield ‘cognitive growth charts’; (ii) develop machine learning-based tools to harmonise disparate cognitive data from independent studies to a common reference to enable this to be applied to a wide range of cognitive instruments; (iii) assemble and curate demographically diverse reference datasets derived from large, population level cohorts in order to define comprehensive reference models that provide equitable predictions for a wide range of sociodemographic, racial and clinical backgrounds. This will yield considerably greater generalisability relative to existing norms for cognitive data. Lastly, we will (iv) use machine learning models on the basis of these models to stratify individuals from early psychosis cohorts and predict their clinical and functional outcomes (e.g. educational and vocational function) across short, medium and long timescales. A crucial feature of this project is that it has been co-developed together with experts with lived experience to promote real-world impact and relevance, which has been shown essential for interpreting findings and shaping research questions.^31^ Guided by the input from these experts we will refine research questions towards concerns they identify using focus groups, dissemination and collaborative governance to ensure that our approach aligns with their perspectives. Ultimately, we hope that providing a nuanced understanding of cognitive impairments in individuals with psychosis can improve individualised care and aligns with the motivations of the lived experience experts contributing to the project.

## Methodology

The PRECOGNITION project comprises partners from the Netherlands (Donders Institute / Radboud University Medical Centre), the UK (Institute of Psychiatry, Psychology and Neuroscience, King’s College London), Norway (Oslo University Hospital, University of Oslo), Denmark (Mental Health Services CPH, Copenhagen University Hospital), and the Czech Republic (National Institute of Mental Health, Klecany). The project is structured around three integrated, yet complementary work packages (WPs), each aiming to solve key challenges (Figure 1).

**Figure 1:**
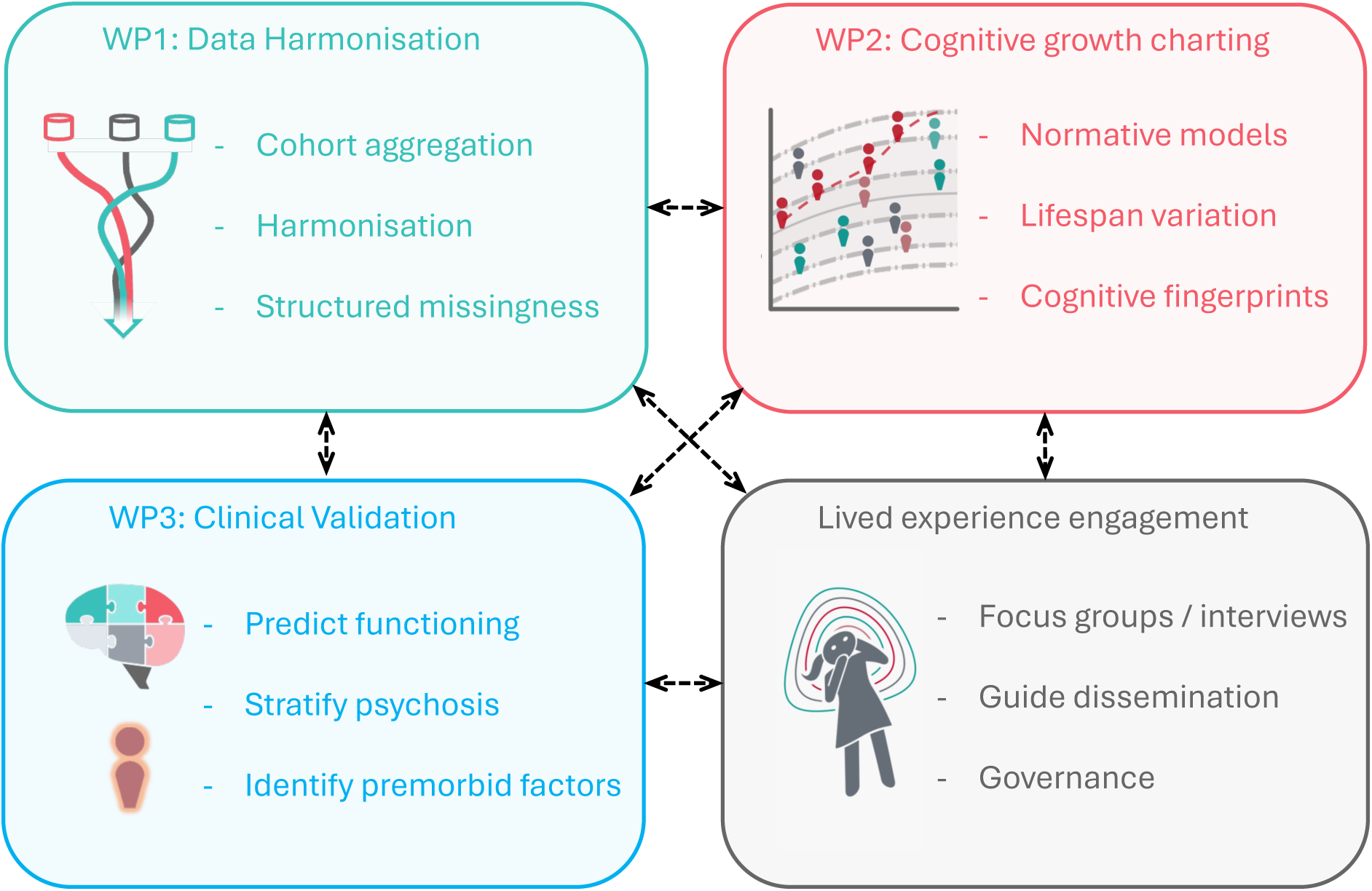
The PRECOGNITION project is structured into three mutually interacting workpackages (WPs), with lived experience engagement throughout the project. Key themes for each WP are identified.

### WP1: Data integration and Harmonisation

Understanding variation amongst individuals with psychosis requires an understanding of how such variation is nested within variation across the wider population and lifespan. This in turn requires large samples and integration across studies to accurately quantify variability across individuals. There are several challenges for this endeavour in psychiatry, the most pertinent are a lack of existing large reference datasets with consistent psychometric instruments and a lack of harmonized assessment across studies. Rather, different studies typically assess cognition using variants of different tests, derived from different cognitive batteries, with poor standardisation across studies. The PRECOGNITION project aims to address this problem and enable multiple datasets to be combined via a novel imputation-based approach to harmonise studies, our proposed high-level framework.^32^ Whilst this approach shares similarities with conventional multiple imputation techniques, it is crucial to recognise that conventional data imputation approaches are unsuitable for this task because of strong dependencies within the pattern of missing data across cohorts, which is referred to as ‘structured missingness’^33^ (see Figure 3 below for an example). Theoretical frameworks and practical tools to accommodate structured missingness are only beginning to be developed within machine learning. Therefore, a key objective of this work package is to develop the appropriate analytical methodology to adequately deal with this task and produce ‘complete’ datasets that allow the estimation of normative models across heterogeneous samples where not all instruments might have been included in each study. This approach will enable us to reconstruct different cognitive instruments and impute missing data whilst faithfully representing the different distributions of different variables in the cognitive battery. This is useful, for example, if certain studies only acquired a subset of tasks from a given cognitive battery. In addition, if the different cognitive batteries measure similar cognitive constructs (e.g. different measures of working memory from different cognitive tasks), this approach will also allow us to translate between instruments that measure the same underlying construct, even if the number of instruments and their distributions differ. We will validate all models extensively, using simulations, out of sample data and validation across and within cohorts.

### WP2: Develop normative models for cognition (‘cognitive growth charting’)

The goal of this work package is to estimate normative models for population-level cognitive data across multiple domains (e.g. intellectual functioning, processing speed, working memory and verbal learning). For example, normative modelling has been shown to be useful for mapping variation in brain imaging measures as a function of clinical covariates, while also accounting for structured variation within the population (e.g. study centre, demographic group, diagnosis).^25,34,35^ and we have also shown proof of concept evidence that it is also suited to cognitive measures.^24^ This allows us to: (i) map centiles of variation across the population that vary smoothly across the lifespan; (ii) make statistical inferences as to where each individual participant can be placed within the population range (e.g. at which centile, and at what level of certainty) and (iii) detect individuals with an atypical or abnormal profile. A key feature of normative modelling techniques is that they enable us to parse heterogeneity at the level of the individual person, without needing to assume that effects overlap across individuals.

It is important to recognise that whilst our approach aligns with the classical motivation for neuropsychological testing in terms of placing individuals within population norms, it also goes considerably beyond conventional statistical approaches for exploiting neuropsychological data for group-level research purposes. In contrast to the conventional approach, which either involves grouping subjects to match particular age bins or, for research purposes, performing a linear regression to adjust for age and then fitting a parametric distribution to the residuals,^36,37^ normative modelling captures the potentially complex shape of different measures, using non-linear centile curves that vary smoothly across the lifespan along with flexible distributions to map variation across individuals and random effect structures to model clinical and demographic structure within the population. This provides multiple benefits in that it: (i) accounts for differences in variance and distributional shape as a function of the input variables and ensures that centiles do not cross across the lifespan;^38^ (ii) allows us to efficiently model different clinical and demographic backgrounds in the same model, for example providing a model that provides norms that are properly adjusted for sociodemographic background and that are calibrated for individuals with psychosis, and which (iii) cleanly separates different sources of variance, for example separating inter-subject variation from uncertainty in the model parameters. This provides more precise statistical inferences and greater power for detecting individual deviations, especially in the outer centiles where the data are frequently the sparsest. Finally, (iv) this approach provides coherent inferences in longitudinal studies with respect to a common reference model, for example by modelling the velocity of cognitive changes across the lifespan.^39^

Since many of the datasets in which we will apply this methodology are longitudinal, we will develop analytical methodology to accommodate longitudinal data, where changes in the expected deviation of subjects across longitudinal timepoints can be detected as centile crossings.^40^ Preliminary data derived from cohorts of individuals with a diagnosis of schizophrenia in neuroimaging give good evidence that normative modelling techniques become exquisitely sensitive for detecting longitudinal changes.^30^ However, we will also develop methodology to model longitudinal change at the individual level, whilst accounting for potentially confounding factors, based on approaches developed within the pediatric growth charting literature.^39,41,42^ This framework will also accommodate potential biases specific to longitudinal cognitive studies, for example those that may be due to practice or motivational effects (e.g. where individuals with a mental illness may have a higher or lower motivation to perform the tests). We will give careful attention to these issues; for example, to differentiate practice effects from clinically relevant variation, we will validate longitudinal trajectories in individuals with a diagnosis of schizophrenia against matched controls.

Finally, we will fit latent variable models (such as structural equation models or canonical correlation analysis) to the individual deviation scores derived from these normative models and use these as a basis for stratifying individuals from psychosis cohorts. These latent variable models –unlike classical clustering methods– do not force individuals to belong to one subtype but rather provide overlapping latent profiles that confer risk or resilience for psychosis, where multiple profiles can be expressed simultaneously within individuals and the same profile can be expressed differently across individuals. We refer to these neuropsychological profiles as ‘cognitive fingerprints’ (i.e. describing the strengths and weaknesses of individuals across different cognitive domains). This approach will provide more precise stratification of individuals and the ability to identify and map the convergence of different biological, environmental and genetic mechanisms on the same clinical phenotype and to identify and utilize complementary information provided by cognition over and above the information provided by neuroimaging and other types of information. We have shown the value of this approach in multiple publications.^43–45^

### WP3: Use neuropsychological profiles to predict outcome and stratify psychosis

In this WP, we will evaluate and comprehensively benchmark the utility of the neuropsychological profiles identified in WP2 to predict functioning in early psychosis, from multiple angles and across short, medium and long timescales. We will also associate the neuropsychological profiles with deviations derived from normative models of neuroimaging features we have already brought online (for example derived from structural MRI and functional connectivity),^46–48^ and with genetic markers. In order to assess functioning broadly, we will begin with symptom measures and measures of overall functioning (e.g. Global Assessment of Functioning scales) derived from clinical follow-up (when available) but we also aim to move beyond these and validate against real-world functional outcomes including educational and vocational status. These outcomes will be co-defined together with the lived experience experts in our team and other consultants. Since this work package is the most extensive and clinically multifaceted, we will subdivide the work according to three clinical objectives:

1. Use cognition to improve prediction of real-world functional outcomes in psychosis
2. Map individual variation in cognition to neurobiology (MRI) to stratify early psychosis
3. Relate premorbid cognition to neuroimaging and genetics using prospective cohorts.

## Cohorts and Measures

To achieve these objectives, we will make use of multiple cohorts, including large population-based cohorts to estimate variation across the spectrum of functioning across different cognitive domains. In order to ensure that the normative models we develop are as broadly representative as possible, we will leverage data from large scale international initiatives including the Human Connectome Project Early Psychosis initiative (humanconnectome.org/study/human-connectome-project-for-early-psychosis) and samples that assess premorbid functioning in healthy cohorts enriched for psychopathology, including psychosis, e.g. the Philadelphia Neurodevelopmental Cohort (PNC)^49^, the Adolescent Brain Cognitive Development (ABCD) study^50,51^ and also the Norwegian Mother and Child Cohort study (MoBa), which is an ongoing population-based prospective birth cohort that follows 114,000 individuals from birth in addition to their fathers and mothers (N>240,000 including both parents).^52^ These cohorts are described in Table 1 below.

**Table 1:** Population based cohorts.

|  | Dataset | Subjects | Ages | Notes |
| --- | --- | --- | --- | --- |
| 1 | ABCD<sup>50</sup> | 11,000 | 9-20 | Longitudinal cohort enriched for risk. With neuroimaging + genetics |
| 2 | Philadelphia Neurodevelopmental Cohort<sup>49</sup> | 10,000 | 5-21 | Neurodevelopmental sample, enriched for risk, neuroimaging in N=1200 |
| 3 | ALSPAC<sup>54</sup> | >10,000 | 18-30 at baseline | Multigenerational longitudinal cohort study with imaging in a subsample |
| 4 | HCP<sup>55</sup> | 1,113 | 22-40 | High-quality neuroimaging data with genetics |
| 5 | HCP Lifespan<sup>55</sup> | 1,000 | 5-90 | High-quality neuroimaging and genetics |
| 6 | IDLaS<sup>56</sup> | 1,000 | 18-60 | Sample with neuroimaging in 200. This sample contains a comprehensive cognitive battery developed at the host institute. |
| 7 | HealthyBrain<sup>57</sup> | 1,000 | 30-40 | Longitudinal population sample with imaging and genetics and the IDLaS battery. |
| 8 | CamCan<sup>58</sup> | 648 | 18-88 | Healthy lifespan sample with extensive phenotyping, neuroimaging and genetics |
| 9 | MoBa<sup>52</sup> | 114,000 (240,000 with parents) | Before birth-age 21 | Ongoing prospective birth cohort of adolescents and their parents with links to school, socioeconomic and Norwegian health registries. |
| 10 | UK Biobank | 500,000 | 50-80 | Population-based cohort of 500,000 participants with genetics, cognition and health data |
| 11 | BRAINMINT<sup>59,60</sup> | 1,200 | 8-45 | Ongoing longitudinal population-based study on brain, cognition and mental health, including MRI, cognitive, genetics and clinical phenotypes with possibility to link to Norwegian population and health registries |

In addition, we will make use of large cohorts of individuals with early psychosis, derived from four European countries. Full details about these samples are provided in Table 1 below. All these samples have extensive cognitive assessments, using standardized tasks plus matched controls, extensive clinical and functional assessments^53^, neuroimaging and genetics.

A unique and compelling feature of our project is that we have access to extensive health, vocational and educational outcomes via Scandinavian registries (for example linked with the MoBa study described above). We will make use of this information to assess functional outcomes as broadly as possible and in a manner that faithfully reflects functioning in the real world, not only over the timeframe of clinical follow-up, but also across the lifespan. Furthermore, as mentioned above, these outcomes will be guided by input from our experts with lived experience.

Another key feature of our project is that most of the cohorts we will use are genotyped and have neuroimaging data in addition to information on environmental stressors allowing us to assess the interplay between cognition, environmental and neurobiological influences.

### Cognitive and outcome measures

Across the different cohorts (summarized in Table 1 and 2), a range of different tests were administered based on a number of standard cognitive batteries including the Wechsler Abbreviated Scale of Intelligence (WASI), Wechsler adult intelligence scale (WAIS),^68^ Brief Assessment of Cognition in Schizophrenia (BACS)^69^, National Adult Reading Test (NART),^70^ Delis-Kaplan Executive Functioning System (D-KEFS),^71^ MATRICS Consensus Cognitive Battery (MCCB),^53^ the Cambridge Neuropsychological test automated battery (CANTAB)^72^ and the National Institute of Health (NIH) Toolbox for the assessment of cognitive function.^73^ Cognitive data for the MoBa sample are provided by the Moba BrainHealth initiative (https://www.fhi.no/studier/moba/undersokelser/hjernehelse/) which invites participants from MoBa to complete online cognitive testing using the Memoro platform (https://memoro.medisin.ntnu.no). BRAINMINT participants have completed a standardized computerized test battery assessing various cognitive domains^60,74^. Note that although the methods we will develop in WP1 will provide item-level predictions, for simplicity, we will group cognitive instruments according to their underlying cognitive domains, namely *intellectual functioning, processing speed, verbal learning and memory, semantic fluency, inhibitory control, working memory and fine-motor speed*.

All clinical studies (Table 2) also have measures of symptom severity including the Positive and Negative Symptom Scales (PANSS) and measures of global functioning including the Global Assessment of Functioning (GAF) scales at baseline and at clinical follow-up. In addition, many of the samples we include allow us to assess functioning in terms of long-term health related outcomes. For example, the UK Biobank has links with primary care health related data and the Scandinavian samples allow us to assess lifetime functioning using health registry outcomes (e.q. quantifying the frequency, timing and duration of interactions with mental health services as well as prescription drugs across the lifespan). More importantly, as outlined above, we are also able to obtain measures of functioning derived, for example, from employment status and other vocational outcomes via the Norwegian Labour and Welfare Administration (NAV) registry (see e.g. https://www.nav.no/en/home/employers/nav-state-register-of-employers-and-employees).

**Table 2:** Psychosis cohorts. Abbreviations: FEP=first episode psychois, SCZ= schizophrenia, UHR=ultra-high risk.

|  | Dataset | Subjects | Ages | Notes |
| --- | --- | --- | --- | --- |
| 1 | TOP<sup>26</sup> (Oslo) | 1,100 SCZ<br>1,200 HC | 20-65 | Cohort study, with imaging and genetics and Norwegian health and population registries. |
| 2 | STRATA<sup>61</sup> (KCL) | 150 FEP | 18-48 | Two sub-studies: 1. Cross-sectional comparison of treatment responders and non-responders. 2. Prospective cohort study of FEP individuals recently starting their first medication, followed for 6 weeks |
| 3 | AESOP<sup>62</sup> (KCL) | 557 FEP | 18-60 | Prospective cohort study, with 10-year follow up, neuroimaging and genetics |
| 4 | GAP<sup>63</sup> (KCL) | 410 FEP<br>370 HC | 18-65 | Cross-sectional study with clinical follow up, neuroimaging and genetics. |
| 5 | Mental Health Services CPH, Copenhagen University Hospital<sup>64-67</sup> | 200 FEP<br>200 HC<br>50 UHR | 18-45 | Cohort studies with longitudinal follow-up and links to Danish registries. This includes sub-studies of medication naïve first episode patients, longitudinal treatment cohorts and ultra-high risk (UHR) individuals |
| 6 | NIMH (Prague) | 200 FEP<br>200 HC |  | Prospective cohort study with longitudinal follow up at 1 year plus imaging. |
| 7 | HCP early psychosis | 320 FEP<br>80 HC | 16-35 | Imaging protocol matched to healthy HCP samples |

We can similarly leverage education registry data (https://www.ssb.no/a/english/mikrodata/datasamling/nudb/nudb_20130607-en.html) to derive proxy measures of educational attainment, as we have done in previous studies.^75^

## Analysis Plan

### Overview

A high-level overview of the analytic workflow we will follow in the project is shown in Figure 2. We follow a similar staged protocol for data harmonisation that we have outlined previously,^32^ which involves the following stages:

**Figure 2:**
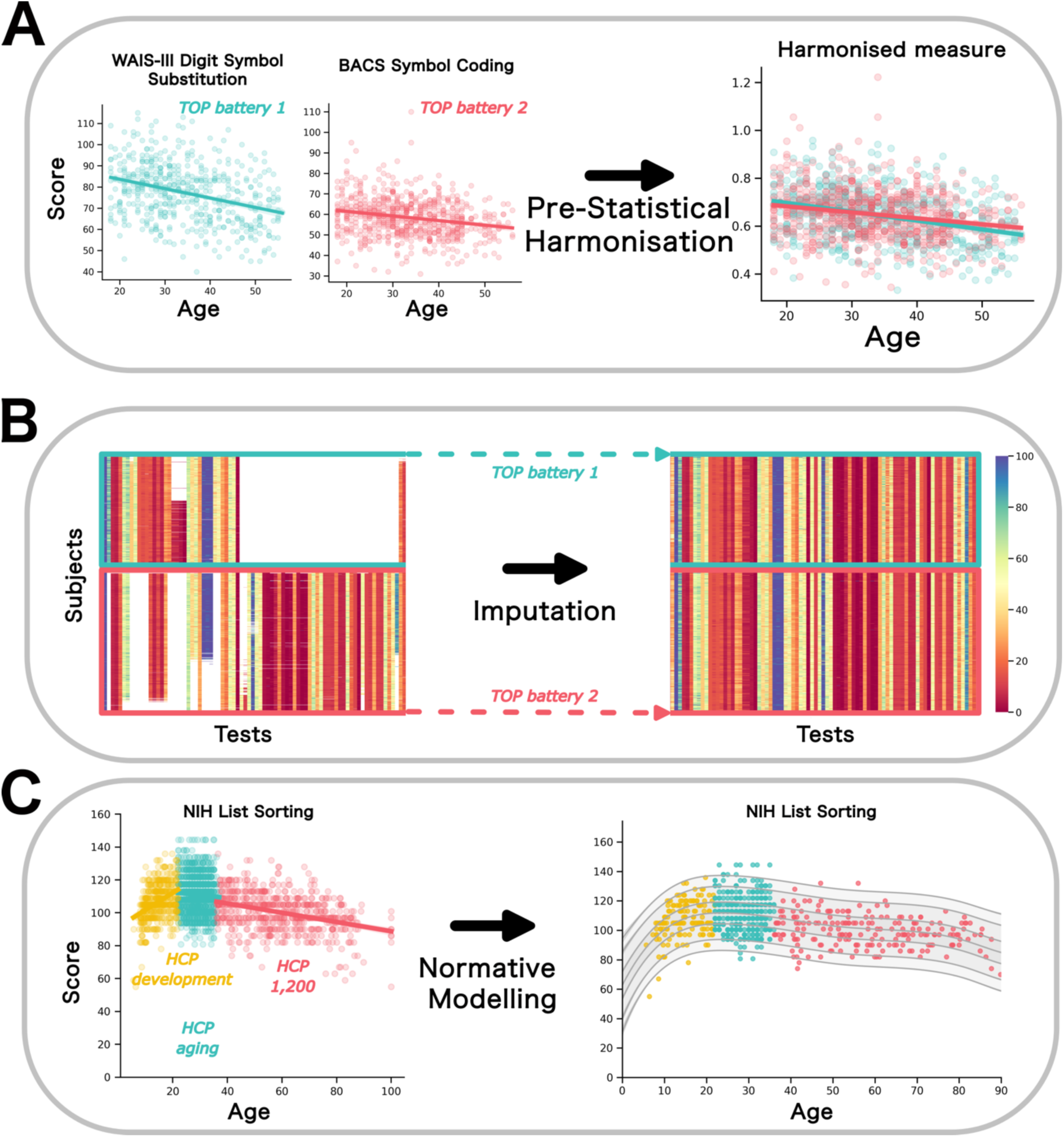
A high-level overview of our analytical workflow including pre-statistical harmonisation (A), imputation (B), followed by fitting normative models (C). In panel A, two measures are shown that are equivalent, but derived from different tests and having slight variations. These can be accommodated by simple mathematical operations such as rescalings. In panel B, we show a graphical representation of data derived from the thematically organised pyschosis (TOP) cohort, which contains two distinct cognitive batteries applied to different subjects, inducing a strong pattern of structured missingness in the data, requiring custom-built imputation techniques to complete (right). In panel C, we show an exmaple of an identical measure derived from the NIH toolbox for multiple studies from the Human Connectome Project lifespan datasets. In this case cohort effects and non-linearity across the lifespan can be accommodated in the normative modelling step.

### Expert review

In this stage, the goal is to determine which instruments can be aligned directly and which instruments assess the same underlying construct, but which may elicit different distributions or have different psychometric properties. This involves input from clinicians, methodologists and psychometricians and the goal is to determine which instruments can be directly aligned by simple operations, which require a more nuanced statistical harmonisation approach, and crucially, which instruments ought not to be combined (e.g. because they measure different underlying psychological constructs).

### Pre-statistical harmonisation

In this stage, data from directly comparable instruments (e.g. different variants of the same test) are combined using simple operations. For example, this might involve rescaling to accommodate structural differences in the tests. A simple example is shown in Figure 2A, where two measures of symbol coding were combined by adjusting for the length of time allowed under the test to perform the symbol coding. As shown in the figure, this yields a measure that has equivalent psychometric properties under the lifespan.

### Statistical harmonisation

In this stage, we will apply an imputation-based approach to combine data from different studies. This is illustrated in Figure 2B, where we show a graphical representation of the cognitive data from the thematically organised psychosis (TOP) dataset which consists of two partially overlapping cognitive batteries that were administered to different subjects (see ^23^). This induces a strong pattern of structured missingness within the data (Figure 2B, left) which will be addressed by custom built imputation techniques to accommodate this structure (right). To achieve this, we will build on the emerging machine learning discipline of structured missingness^33^ which involves building a generative model (e.g. implemented using Bayesian hierarchical models) that aims to model the missing data mechanisms as accurately as possible during the imputation process.

### Normative modelling

Finally, we will estimate normative models for each harmonised measure. In order to estimate the normative models, we will employ hierarchal Bayesian regression techniques that we have developed for neuroimaging.^34,35,76^ These provide several important features for our purposes, namely sufficient flexibility to model site effects, non-Gaussianity and heteroskedasticity. This will also allow us to model clinical variables as random effects, effectively allowing us – for example – to estimate reference models that provide distinct centile curves for individuals with and without psychosis, also accounting for sociodemographic characteristics. We will extensively validate all models out of samples out of sample and across cohorts to ensure generalisability. An example is shown in Figure 2C that illustrates a cognitive measure derived from the NIH toolbox that shows non-linearity across the lifespan.

### Application to clinical cohorts

We will then apply these models to the clinical cohorts in order to generate deviation scores for individuals with psychosis, which will be validated extensively within and across cohorts. We will then apply latent variable models to the deviation scores in order to obtain latent profiles across cognitive domains, for example Bayesian non-parametric variants of factor analysis that properly accommodate the distribution of different measures.^77^ Next, we will determine whether these predict both clinical and functional outcome measures using penalised regression models and against external measures (for example imaging and genetics), which can provide additional validation to support the stratifications we will derive. Finally, we will evaluate different proxy measures for cognition(e.g. secondary school educational attainment), that can be applied to large scale studies, and then use these to search for pre-morbid markers in prospective population cohorts with data acquired before the onset of the first psychosis episode (for example MoBa^52^).

## Preliminary Results

We show examples of data derived from existing studies in Figure 3, which illustrates several of the key problems this project aims to solve. In the left panel, we plot the scores from the California Verbal Learning Test (CVLT)^78^ and the Hopkins Verbal Learning Test (HVLT)^79^ as a function of age. This shows two challenges: first, the CVLT shows clear age-related heteroskedasticity in that the variance increases with age. This should be accounted for in the modelling in order to ensure accurate inferences. Second, these tests should – from a theoretical perspective – measure the same underlying construct (i.e. verbal learning), but the CVLT involves recalling a list of 15 words, relative to 12 words from the HVLT. While it may be tempting to combine these instruments it is clear that this change to the test also changes its psychometric properties over the lifespan in that the CVLT shows age-dependence, whereas the HVLT does not. In the middle panel, we show an instrument, the ‘Mazes’ score from the MATRICS battery (assessing planning and foresight), that was administered during the second TOP battery, but which does not have a direct correspondent in the first TOP battery (although other tasks measures related abilities). This therefore needs to be addressed via a statistical approach. In the right panel, we show reading age and picture vocabulary scores derived from the NIH toolbox^73^ which shows: (i) highly nonlinear effects across the lifespan and (ii) strong cohort effects, despite the same test being administered across different cohorts. Again, these effects must be properly accounted for in the modelling.

**Figure 3:**
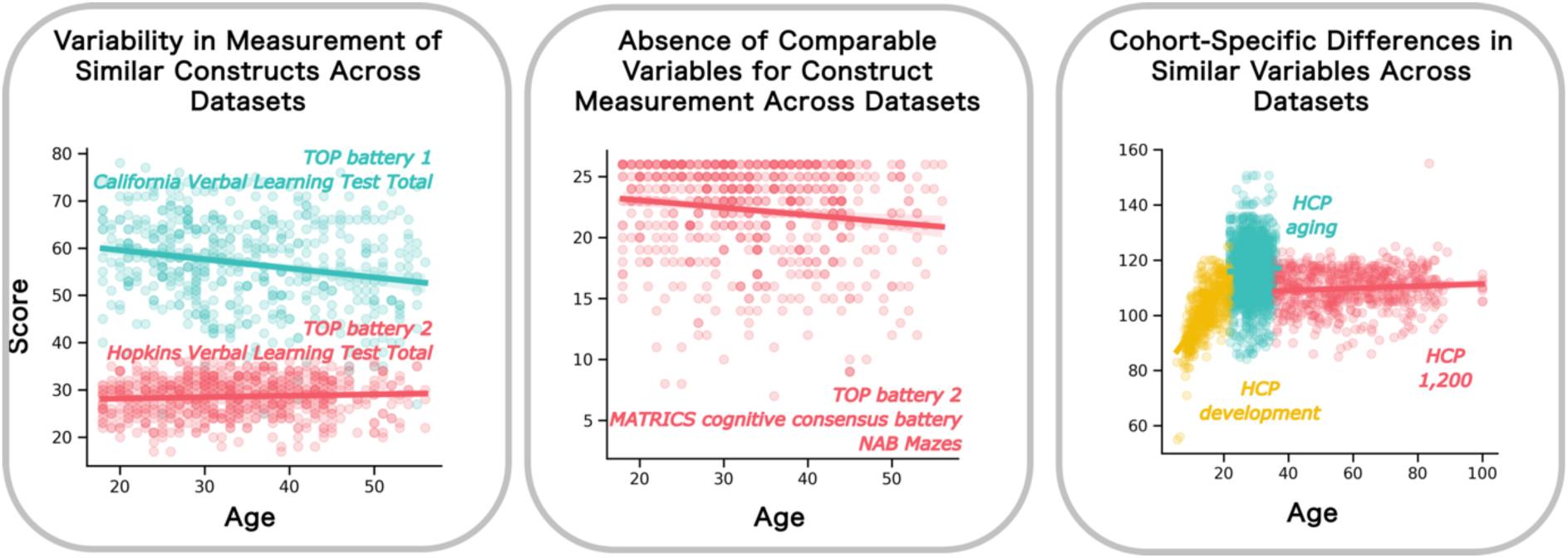
Illustrative data assembled from some of the cohorts used in the PRECOGNITION project. Left panel: Delayed recall scores from the California Verbal Learning Test and Hopkins Verbal Learning Test, derived from the different Thematically Organised Psychosis (TOP) batteries. Although the tests are nearly identical, they have different psychometric properties across the lifespan. Middle panel: MATRICS ‘Mazes’ score (assessing planning and foresight) used in the second TOP battery shows clear heteroskedasticity across the lifespan and does not have an equivalent in the first TOP battery. This means it must be reconstructed using imputation. Right panel: Reading age score from the Human Connectome project (HCP) lifespan data. This shows a non-linear relationship with age and strong cohort effects, which must be accommodated during the modelling.

## Lived Experience Involvement

A notable feature of the PRECOGNITION project is engagement throughout with lived experience experts to guide the project throughout its duration. This input is multi-faceted but especially important topics include ensuring that the project focusses on functional outcomes most relevant to individuals with psychosis and discussing the broader societal implications of the project outcomes from the outset. These activities will be coordinated with involvement of lived experience experts within the project team, their network of people with lived experience of psychosis in the UK, in Norway and more broadly through EU Networks we are part of (e.g. the <u>European Brain Research Area</u> and <u>GAMIAN-Europe</u>). At the outset of the project we conducted a launch event attended by scientists and individuals with lived experience in order to gain lived experience feedback.

Early in the project, we will conduct focus groups and interviews with individuals with lived experience to provide us background information in order for us to define, select and refine functional, clinical and psychosocial outcomes for us to focus on as primary endpoints. These will be conducted by the study team, guided by input from members of the project team with lived experience of psychosis who have guided the selection of questions to include. More specifically, at study onset, we will conduct one-to-one interviews with people with lived experience of psychosis and focus groups with unpaid carers of people with lived experience of psychosis at each of the main clinical sites (London and Oslo). During the one-to-one interviews, we will aim to understand what are the cognitive deficits that individuals with psychosis consider to be the most impairing. During the focus groups, we will discuss the carers’ personal experience of caring for a person with psychosis and cognitive impairments. This will help the study team decide which to consider as endpoints for the prediction models we will develop in WP3. Focus groups and one-to-one interviews data will be transcribed verbatim and subjected to reflexive thematic analysis.^80^ Reflexive thematic analysis is a theoretically flexible approach widely used in mental health research to analyze qualitative data.^81–83^ Reflexive thematic analysis will include inductive and deductive coding, to ensure that the analysis is shaped by participants’ priorities and concerns.^84^ This will allow the study team to systematically probe the data, asking questions arising from the existing literature. At the end of the project, we will organise seminars and webinars with people with lived experience of psychosis to present the results of the models derived in WPs 1 and 2, discuss risks and benefits of using these predictive models, and include recommendations regarding future stages of research. host webinars to communicate our findings to the lay public, healthy personnel and people with lived experience. Again, we will conduct these activities in tandem with lived experience experts.

In addition, we will rely on lived experience experts within our project team and external lived experience experts to help us with other tasks, including discussing and interpreting study findings and in providing input into articles specifically directed at mental health organisations to describe our research. Finally, we will rely on our lived experience experts to guide the dissemination of our results, to ensure that the communication of our findings at scientific fora, to service user communities and to the lay public is, clear, understandable, ethically informed and sensitive to the potential issues of inadvertent stigmatization of groups of individuals. As noted above, an important aspect of the work will include clarifying the ethical and user relevant aspects of applying prediction models in psychiatry, as well as clarifying the clinical and societal aspects of the project outcomes.

## Conclusions and Expected Outcomes

We have outlined the rationale, design and analysis plan for the PRECOGNITION project, which aims to develop cognitive growth charting approaches for psychosis. The PRECOGNITION project will not generate any new primary data, but by leveraging existing data in novel ways, it will produce multiple distinct contributions including (i) guidance informed by lived experience as to functional outcomes that can inform this and future studies; (ii) novel methodological approaches for normative modelling of large scale cognitive data, including approaches for longitudinal tracking and prediction at the individual level and for combining data from different tasks that measure the same underlying construct (iii) machine learning models for stratifying cohorts on the basis of these models; (iv) comprehensive reference models that provide broad sociodemographic coverage including different languages and distinct norms for individuals with psychosis and unaffected individuals. We intend that these models will supersede current norms currently used in the field^37^ and also provide domain specific norms; (v) software tools to enable these that we will distribute via our online platform^48^ and finally (vi) stratification models and predictions for functional outcome measures in psychosis cohorts. If successful, these models could be widely deployed, for example via mobile or tablet technology. This will provide a platform to maximise the value of cognitive instruments in predicting the course and functional outcome of psychosis.

## Data Availability

No primary data will be gathered during this study

## Acknowledgments

This study was funded by the Wellcome Trust under grant number 226706/Z/22/Z

